# Saccade dynamics in the recovery phase of abducens nerve palsy

**DOI:** 10.1101/2021.11.29.21267015

**Authors:** E Kemanetzoglou, KI Chatzistefanou, N Smyrnis, E Kararizou, E Anagnostou

## Abstract

**INTRODUCTION:** Despite the fact that abducens nerve palsy (ANP) is the most common ocular motor palsy, the literature on the respective saccade dynamics, both in the paretic (PE) and non-paretic eye (nPE), is scarce.

**AIMS AND METHODOLOGY:** The aim of this study was to examine the maximum velocity, duration and accuracy of horizontal saccades, in individuals with unilateral ANP, and to compare them with normal controls. Binocular horizontal eye movements were recorded at 5°, 10° and 15°, using an infrared corneal reflection device from 21 adults with microvascular unilateral ANP during the acute and the chronic phase of the palsy, as well as 18 healthy adults. Non-parametric tests were used for statistical comparisons.

**RESULTS:** The PE, when compared to the nPE, presents a slightly lower saccadic amplitude and velocity/amplitude ratio and a higher duration/amplitude ratio. The nPE, compared to the healthy eye (HE) of the control group, showed consistently amplitude gain >1 while the velocity/amplitude ratio did not differ in either session. The duration/amplitude ratio tended to be higher in the nPE. The prism dioptres of the PE did not appear to correlate with any parameter tested (amplitude gain, velocity/amplitude ratio, duration/amplitude ratio) of the open nPE, but the amplitude ratio was statistically lower during the first session when the nPE was kept covered and the duration/amplitude ratio decreased significantly.

**CONCLUSIONS:** One of the main findings of the study is the increase in saccade duration during adaptation of ANP. Specifically, the nPE performed orthometric saccades with a longer duration than healthy controls. Given that the motor command reaches the ocular muscles by neural discharges with a “pulse-step” pattern, any adaptation reflects in a change of this pattern. Cerebellar learning leads to an increase in the pulse width of the neural discharge. This idiosyncratic response may be related to plastic changes in central structures that serve learning processes such as the cerebellum. Further research could provide more insight into the cerebellar plastic processes involved in the saccadic adaptation.

## INTRODUCTION

In an ever-changing visual environment, eye movements during fixation and scanning are extremely important for visual world perception. The optimal ocular motor response to the appearance of a target of interest in the periphery of our field of vision is an ocular movement that quickly and accurately leads the target to the macula, something that can be achieved with saccades (1). Most ocular movements in everyday life are saccadic, alternating with fixation, allowing thus the scanning of the environment, in order to maintain the object of interest on the fovea, the retinal area with the highest visual acuity, despite possible movement of the observer or the object. The saccadic movements are ballistic, short, voluntary, conjugate eye movements, which abruptly change the focus and align the fovea with the object of interest. Saccades change the gaze from one target to another, while multiple neural circuits are involved in their control (2).

Abducens nerve palsy (ANP) is the most common ocular motor paralysis, with horizontal diplopia in binocular vision, in the direction of action of the paretic muscle (respective lateral rectus) especially in more distant targets (3). ANP in adults is more often associated with vascular disease and usually a complete or partial improvement is observed after months (4–6).

Saccades are the most studied ocular movements (7) since they constitute, an important diagnostic tool for the control of ocular motility in various diseases (8–10) and an exceptional system for the research of neural motor control. However, literature is scarce about the effects of unilateral ANP on eye movement dynamics during saccades and the variation of the endpoint of the PE and the nPE (8,11–16). Consequently, the saccade adaptation in binocular vision during ocular palsy is still under investigation. In more practical terms, evaluation of saccadic movements in ANP can be an extremely useful clinical tool in locating the lesion and determining the diagnostic approach, without subjecting the patient to expensive and time-consuming diagnostic tests (8,17–19).

## AIM OF THE STUDY

The aim of the study was to investigate and record the saccade dynamics (maximum velocity and duration) and accuracy of the horizontal saccades, in individuals with unilateral ANP, during the acute and chronic phase of the palsy. We also aimed to delineate differences in saccade characteristics between the patients with ANP and an age-matched control group.

## METHODOLOGY

### PARTICIPANTS

A sample of twenty-one adults with microvascular ANP were included in the study. All patients experienced diplopia and presented with a compensatory head turn. The “microvascular” etiology of ANP was based on: the presence of diabetes mellitus or high blood pressure, a negative brain imaging (MRI or CT scan), spontaneous palsy improvement no later than three months after the onset of symptoms and complete palsy recovery no later than six months after the onset of symptoms. Patients had to meet all four criteria in order to be included in the patient group. Pain in the first days or for some time before the onset of the nerve palsy was considered a supportive but not necessary criterion (20). Patients with severe ANP with an extreme axis deviation due to an excessive tonic esotropia, whose palsied eye could not exceed the midline during attempted abduction were not included in this study. The control group was composed of eighteen, age-matched healthy adults who underwent the same tests. All subjects gave written informed consent for participation in the study which was conducted in adherence to the principles of the Declaration of Helsinki and was approved by the Ethics Committee of the First Department of Neurology of the National and Kapodistrian University of Athens.

### RECORDING DEVICE

The study was performed in the Neuro-ophthalmology and Balance Research Laboratory of the First Department of Neurology of the National and Kapodistrian University of Athens at the Aeginiteion Hospital. Binocular horizontal saccades were recorded with an infrared corneal reflection device (IRIS-Skalar, Delft-Netherlands). This system tracts the reflection of infrared radiation of the limbus. Eye position signals were low-pass filtered with a cutoff frequency of 70 Hz, smoothed with a Savtizky Golay filter (2^nd^ degree/8 samples) and digitized with a 14-bit analogue-to digital-converter, sampled at 500 Hz with a National Instruments external card. The digital signal is transmitted to a computer software developed in a Labview environment (NI, Austin, TX, USA). The digitized time series concerning the position of the right and left eyes are then further processed with software developed specifically for the needs of the present study in Matlab code (The Mathworks, Natick, MA).

### MEASUREMENT PROCEDURE

Horizontal rightward and leftward saccades were recorded binocularly, while subjects looked at LED targets placed at ±5°, ±10° and ±15° eccentricities. In the present study, all saccades were made voluntarily between consistently present and lit pair of Led targets throughout the session, in contrast to other ocular motor studies using targets that flash abruptly by releasing reflective saccades. This process best mimics natural ocular motor behavior, where in realistic conditions, people constantly perform saccadic movements on objects that pre-exist in the visual scene that surrounds them by scanning space and refocusing on objects of interest [Figure 1] (21,22).

**Figure 1.**
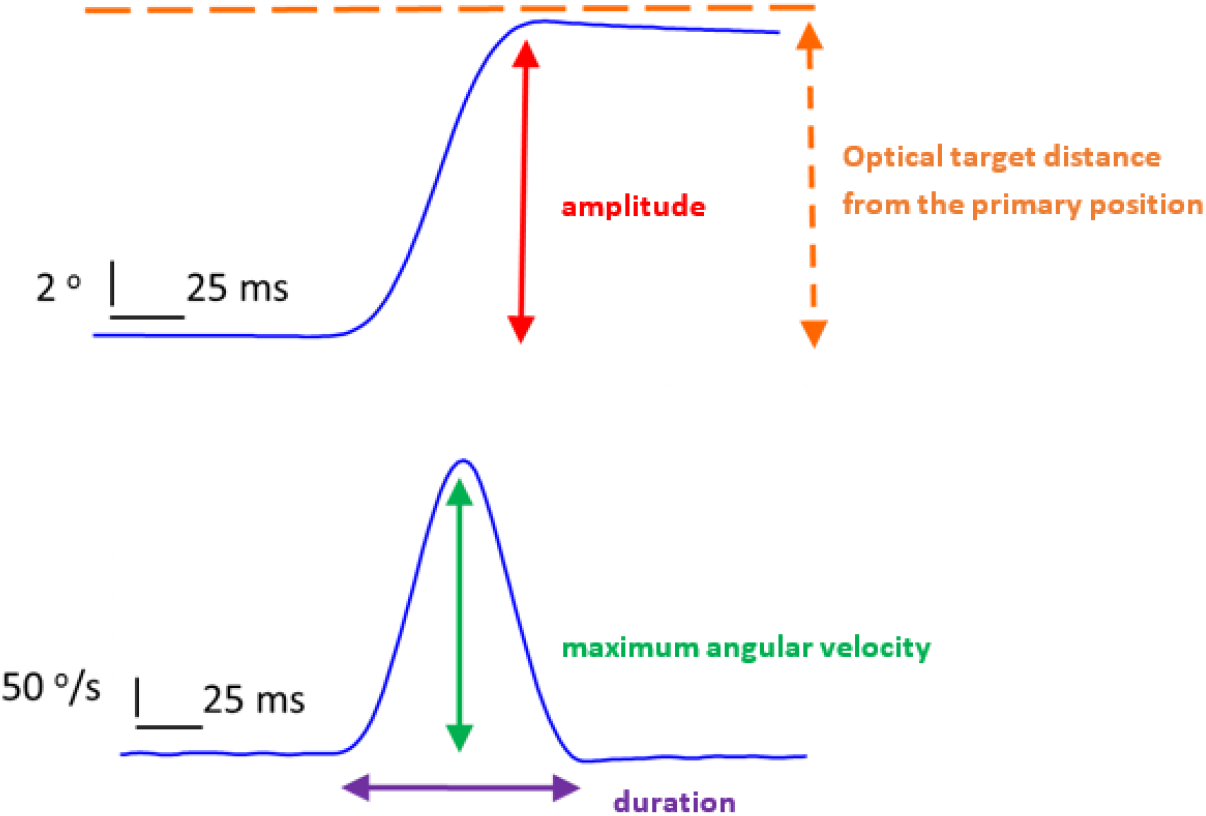
Example of a horizontal rightward 10°, visually guided saccade recorded from a normal control subject with the eye open. The dynamic parameters appear both in the position (upper diagram) and in the velocity profile (lower diagram) of the eye respectively. The horizontal axis gives the time and the vertical the position (upper diagram) or velocity (lower diagram).

During the recordings, the subjects were seated in a dimly illuminated room, stabilizing their head position by using a chin rest. The calibration of the recording device was performed monocularly, while the other eye remained covered: subjects were asked to fixate back and forth between a central and two peripheral LED light dots located at 10° (or 5° when the lateral rectus muscle palsy was severe) on either side from the primary position of gaze, performing a series of horizontal saccades. Then, while one of the two eyes remained covered, in order to prevent fusion, the targets were placed 5° on either side of the primary position of gaze. The saccade recording started and the subject performed 5 rightward and 5 leftward saccades from the central fixation point. The same process was repeated after positioning the targets at 10° and then at 15°. The eye cover permitted movement recordings of the non-viewing contralateral eye. Subsequently the cover was moved to the contralateral eye and the whole procedure was performed from the beginning. Saccade movement data were collected from the fixating (uncovered) paretic eye [PE], from the covered PE, from the fixating (uncovered) non-paretic eye (nPE) and from the covered nPE in the patient group. In the control group, data were collected from the fixating (uncovered) eye (HE) and the non-fixating eye, and after reversing the eye-cover, data were collected again.

### STATISTICAL ANALYSIS

The demographic data of the participants (age, sex) as well as clinical data concerning the group of patients (medical condition, prism diopters) were summarized by descriptive statistical methods (Tables 2 and 3).

The mean age of patients was 60 years (range 20 - 80 years), 57% were male and 52% of patients had left eye ANP. The mean time from the onset of palsy to the first recording session was 30 days (range 2 - 82 days). In the first recording session the average of prism dioptres was 25^Δ^ (range 5^Δ^ – 40^Δ^), while in the second the average was 0 (range 0 – 10^Δ^). The average time period in days between the first and second recording session was 85 days (range 24 - 180 days). In the control group 61% of the subjects were female (39% male) and in 78% of them the dominant eye was the right (22% the left), with average age 58 years (range 34 - 77 years).

The saccade dynamics used for analysis were:

1. The amplitude gain which is the amplitude of the saccade to its desired amplitude ratio (amplitude/desired amplitude) (ie the distance of the optical target from the center)
2. The peak velocity-to-amplitude ratio, which derives by dividing the maximum saccade velocity to the saccade amplitude (velocity/amplitude)
3. The duration-to-amplitude ratio which corresponds to the duration/amplitude fraction of the saccade

The aforementioned dynamics concerned only the primary saccadic movement performed by the subject. The following, smaller, corrective saccades that followed the primary saccade in some cases, were not included in the analysis.

The data of the amplitude gain, the maximum velocity/amplitude ratio and the duration/amplitude ratio, deviated significantly from the Gaussian distribution according to the Kolmogorov-Smirnov test applied to each variable separately. Thus, non-parametric tests were used for the statistical comparisons. The SPSS software version 21.0 (IBM, Armonk, New York) was used for the statistical analysis with the level of significance set at 0.05.

## RESULTS

### COMPARISON OF THE HEALTHY EYES (HE) OF THE CONTROL GROUP WITH THE NON-PARETIC EYE (nPE) OF THE PATIENT GROUP

Since the final ocular motor pathway (abducens nerve - neuromuscular contraction - lateral rectus muscle) is considered evenly intact in both groups, any differences in saccade dynamics must be attributed to central nervous system (CNS) adaptive changes. The movement in the field of action of the palsied lateral rectus muscle during the acute and the recovery phase was compared with the respective movement of the HE of the control group. The recordings were made first with the nPE open and then covered (and the PE covered and open respectively). Given the non-normal distribution of the vast majority of the drawn data, the central tendency and dispersion of the data are displayed as median and 25th/75th percentile respectively, while comparisons between HE and nPE were made by the Mann-Whitney test, a non-parametric test for independent values.

When the HE and the nPE was compared and the nPE was either open or covered, the amplitude gain was fixed > 1 for all optical targets (5°, 10° and 15°). In some cases, in fact, the 75th percentile of the distribution displayed values close to 1.5 when the nPE was open and close to 2 when it was covered, indicating that the saccade was 50% or 100% the size of the original retinal error. This did not assume statistical significance in either of the three optical target distances and in either of the two recording sessions, with the exception of the amplitude gain of the nPE, when it was kept covered, which was statistically higher than that of the control group at 15° [1.18 (0.99-1.42), ρ <0.05]. No correlation was noted between the two groups regarding the velocity/amplitude ratio in any of the conditions in either session. Similarly, the duration/amplitude ratio did not differ between the two groups. There is, however, a trend for higher values in the nPEs of the patient group, especially with regard to more eccentric saccades (10° and 15°) but this assumed statistical significance only at 15° saccades during the second recording session when the nPE was covered [7.0 (5.5-8.7), ρ <0.05].

### CORRELATION BETWEEN CLINICAL DEFICIT OF PATIENTS AND PERFORMANCE OF NON-PARETIC EYE (nPE)

In order to examine whether the angle of deviation is correlated with saccade dynamics of the nPE, the linear correlation coefficient “ρ” was calculated by the non-parametric Spearman Correlation. The velocity/amplitude ratio of the nPE was not significantly correlated with the prism dioptres of the PE in either session regardless whether the nPE was fixating or remained covered. Likewise, the amplitude gain and the duration/amplitude ratio of the open nPE was not correlated, in either session, with the prism dioptres of the esotropia of the PE, but when the nPE was kept covered during the first recording session, the greater the degree of esotropia of the PE in diopters, the lower the amplitude gain was noticed statistically (r=0.4.76, P<0.01). The duration/amplitude ratio was significantly reduced during the first recording session assuming statistical significance in the condition when the nPE was covered, according to the esotropia prism dioptres of the PE (r=-0.404, ρ<0.05). These correlations were absent during the second recording session.

### COMPARISON OF PARETIC (PE) WITH NON-PARETIC EYE (nPE) (PATIENT GROUP)

The non-parametric Wilcoxon signed-rank test was used in order to draw comparisons between the PE and the nPE, during attempted conjugate eye abduction, in the field of the palsied lateral rectus muscle. As expected, with the PE open, the amplitude gain was slightly reduced, but close to 1, in every optical target recorded (5°, 10° and 15°). In the first session, however, it was significantly lower than the corresponding nPE amplitude gain, and this was statistically significant at the more eccentric optical target distances (10° and 15°) [for 10° PE 0.77 (0.67-0.86), nPE 1.08, (0.86-1.50), ρ <0.05] [for 15° PE 0.79, (0.66-0.82), nPE 1.10 (0.92-2.29), ρ <0.01]. In the second session, despite the tendency of slightly lower gains in the PE, this did not assume statistical significance. The PE amplitude was reduced in all three target distances (5°, 10° and 15°), both in the first and, to a lesser extent, in the second recording session, when the PE remained covered [5°, 1^st^ recording session, PE 0.83 (0.58-0.94), nPE 1.02 (0.97-1.16), ρ <0.01, second recording session PE 0.88 (0.75-1.05), nPE 1,04 (0,88-1,13), ρ <0.05] [10°, 1^st^ recording session 10° PE 0.71 (0.53-0.86), nPE 1.33 (1.02-1.43), ρ <0.001, 2^nd^ recording session 10° PE 0.92 (0.82-1.00), nPE 1,06 (0.94-1.20), ρ <0.05] [15° 1^st^ recording session PE 0.65 (0.55-0.86), nPE 1.22 (1.07-1.72), ρ <0.01, 2^nd^ recording session PE 0.93 (0.85-1.01), nPE 1.17 (0.95-1.38), ρ <0.05]. These obvious differences are due to both the expected low range of motion of the PE and the partially hypermetric range of motion of the nPE.

With either eye open, the velocity/amplitude ratio was slightly lower at all target distances (5°, 10° and 15°) in the PE compared to the nPE in both the first and second recording sessions although it assumed statistical significance, only at the 5° in the second session when the PE was open [5° PE 27.1 (23.9-29.7), nPE 30.9 (26.6-37.0), ρ <0.01] and in the first recording session when the nPE was open [5° PE 26,6 (20,8-29,6), nPE 30,4 (26,7-32,9), ρ<0.05].

When the PE was fixating, the duration/amplitude ratio was generally higher in the PE compared to the nPE. This assumed statistical significance in the first session and specifically at 5° and 10°, [5° 1^st^ session PE 18,3 (14,7-25,2), nPE 13,7 (11,3-17,0), ρ<0.05, 10° 1^st^ session PE 12,6 (9,2-14,0), nPE 9,1 (6,8-11,3), ρ <0.05] [Figure 2]. When the fixating eye was the nPE, the longer duration of the saccades of the PE was distinct, since the differences are statistically significant at 5° and 10° during the first, and at 5° during the second recording session [5° 1^st^ session PE 17.1 (14.7-27.4), nPE 13.4. (12.2-16.8), ρ<0.01, 2^nd^ recording session PE 16.5 (12.7-19.3), nPE 13.8 (11.1-15.0), ρ <0.05] [10° 1^st^ session PE 11.5 (9.3-16.5), nPE 9.5 (7.9-11.1), ρ <0.05] [Figure 3 & 4].

**Figure 2:**
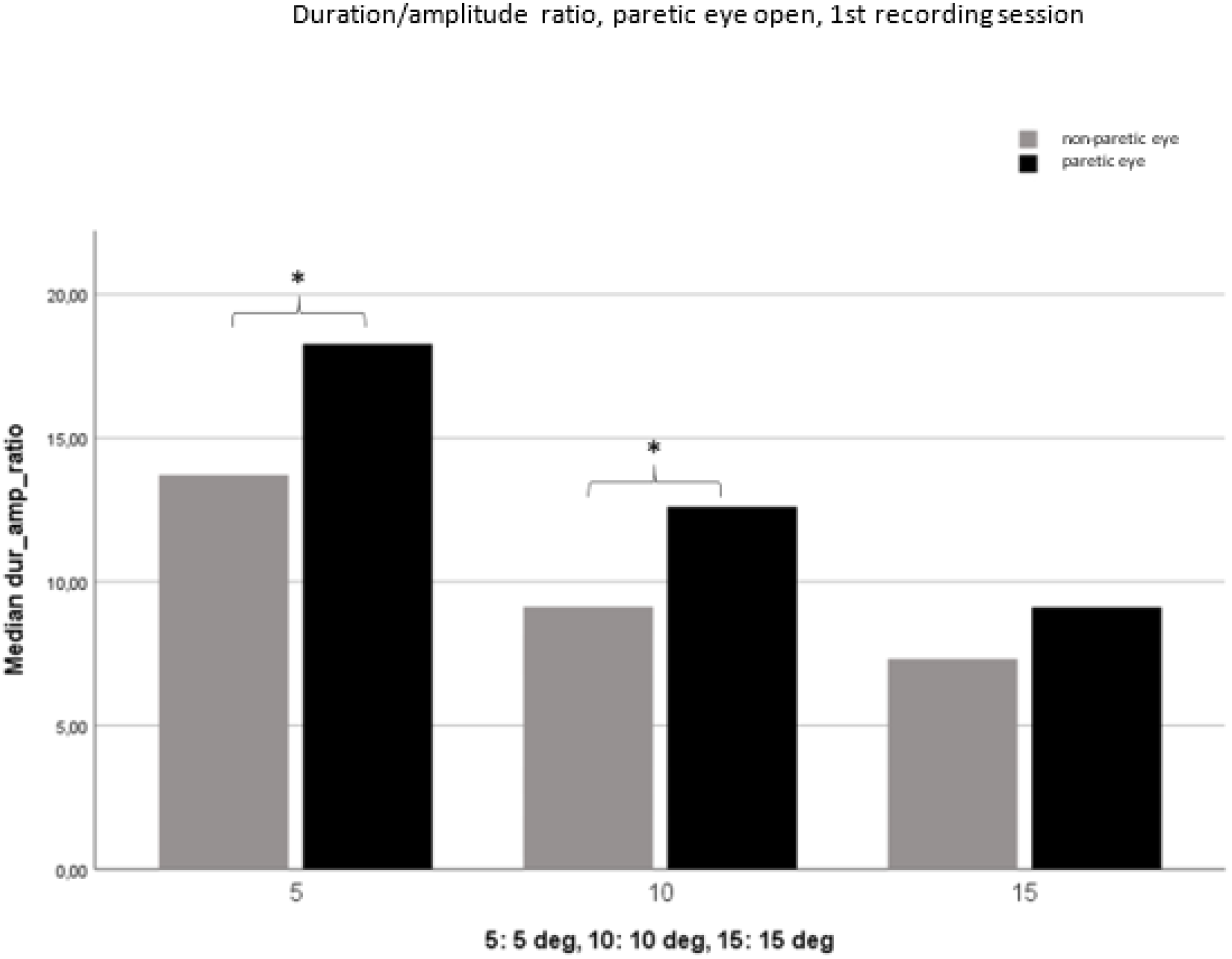
Duration/amplitude ratio with the paretic eye open. Comparison of paretic with non-paretic eye (1st recording session). Significant differences are seen between 5° and 10° (P <0.05, *).

**Figure 3:**
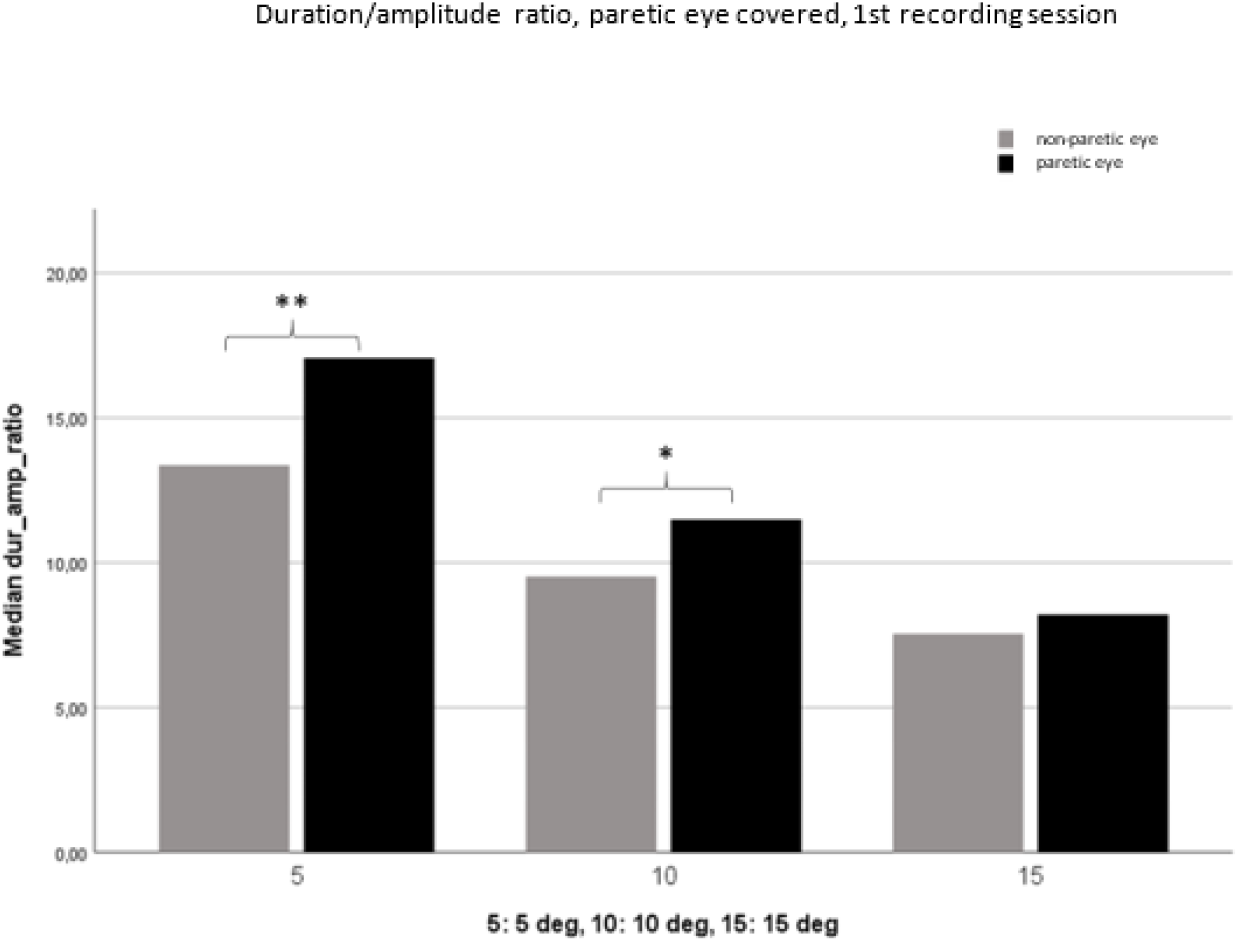
Duration/amplitude ratio with the PE covered. Comparison of paretic with nPE (1st recording session). Statistically significant differences are found at 5° (P <0.01, **) and 10° (P <0.05, *).

**Figure 4:**
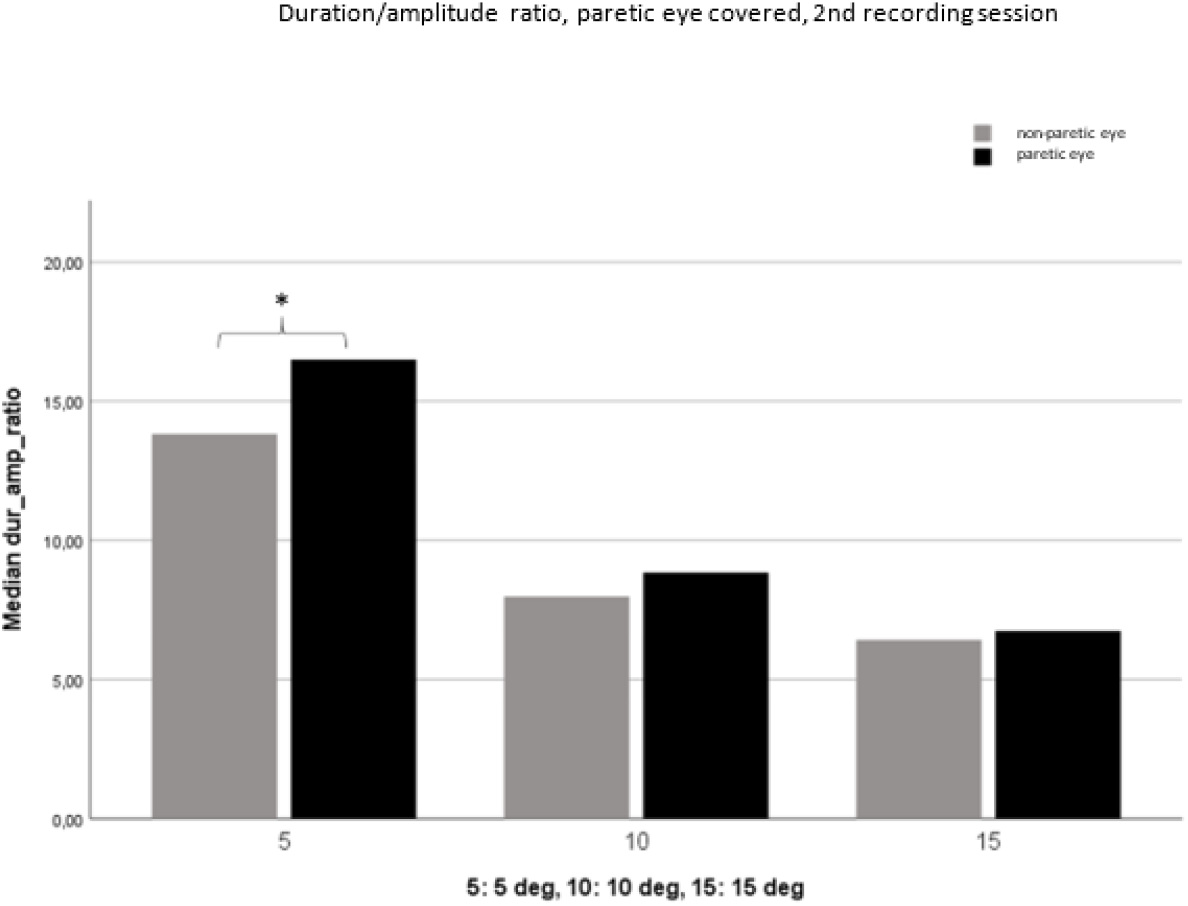
Duration / amplitude ratio with the PE covered. Comparison of paretic with nPE (2^nd^ recording session). The difference of 5° is statistically significant (P<0.05, *).

## DISCUSSION

### AMPLITUDE GAIN

When the PE was fixating (and at the same time the nPE covered), the amplitude gain was slightly reduced, but close to 1. However, at the first recording session, it was significantly lower than the respective amplitude gain of the nPE, and this assumed statistical significance at more eccentric optical targets (10° and 15°). In the second recording session, the saccades continued to be slightly more hypometric compared to those of the nPE, though no longer significant. However, when the patient used the nPE to refocus his gaze (the PE covered), the PE showed hypometric saccades, even at 5°. These differences were observed again, albeit to a lesser extent, in the second recording session. The importance of the retinal error (the distance between primary saccade endpoint and optical target at the macula), becomes clear, so that the saccades of the PE have the correct amplitude in both the acute phase and the recovery phase. With the PE covered, the absence of retinal error (since the nPE focuses correctly) leads to a low range of saccadic movements of the PE. In contrast, the appearance of retinal error when the experimental condition forces the PE to focus on the target, leads to an improvement in its saccadic performance. It is also interesting to note that subjective correction of the patient’s diplopia can be achieved by improving the saccadic dynamics over a period of months, without necessarily achieving a complete objective restoration of the range of saccadic movements when the nPE is used for fixation. According to Hering’s law, any motor command changes of the PE are also expected to manifest in the saccadic behavior of the nPE due to linked innervation. Therefore, once more, the retinal error present during the attempted fixation of the PE appears to lead to an amplification of the central motor command, and, according to Hering’s law, to an increased range of motion of the nPE compared to the completely HE.

### VELOCITY-TO-AMPLITUDE RATIO

The PE displayed a slightly lower velocity/amplitude ratio compared with the nPE in both the first and second recording sessions was evident, regardless whether it remained open or was covered. This difference assumed statistical significance, only at 5° in the second recording session when the PE was open, and in the first session when the PE was kept covered. The velocity/amplitude ratio was largely the same in patients and healthy controls when the HE of the controls and the nPE of the patients were compared in either recording session.

### DURATION-TO-AMPLITUDE RATIO

With the PE open, the duration/amplitude ratio is generally higher in the PE compared to the nPE. This assumes statistical significance in the first session and specifically at 5° and 10°, while at 15° the difference is statistically insignificant. The latter also applies to the small differences observed in the second recording session. The increased duration of the saccades becomes more pronounced when the PE is covered, specifically the differences compared to the nPE were statistically significant at 5° and 10° during the first, and at 5° during the second recording session.

Although the nPE tended to have shorter saccadic durations compared to the PE, as previously mentioned, its comparison with the HE of the control group revealed an interesting result. Initially, with the nPE open, the ratio duration/amplitude did not differ between patients and healthy controls. However, the tendency for higher values in the nPEs of the patient group became apparent, especially with regard to the more eccentric saccades (10° and 15°), though it was not statistically significant. When the nPE was covered, and therefore the ocular motor system was faced with varying degrees of retinal error due to the attempted fixation of the PE, the duration/amplitude ratio showed the same tendency in both the patient and the control group: higher values in the nPEs of the patient group, mainly regarding the more eccentric saccades (10° and 15°). Once more, however, not statistically significant, with the exception of the duration/amplitude ratio during the second session and only for the most eccentric optical target (15°).

### CORRELATION OF NON-PARETIC EYE (nPE) PERFORMANCE WITH CLINICAL DEFICIT

When the nPE was fixating, the amplitude gain did not show any linear correlation with the prism dioptres of the clinical esotropia. When the nPE was kept covered during the first recording session, the amplitude gain was shown to be statistically reduced inversely proportional to the degree of esotropia of the PE measured in dioptres. No statistically significant correlations emerged in the second recording session. It seems, therefore, that the increase in the amplitude of the nPE, which should be perceived as an increased effort of the central ocular motor system to compensate for the motor deficit of the PE, is not greater than the initial clinical deficit, which indicates that the compensatory excess of central command varies from person to person. This idiosyncratic ability to escalate muscle innervation seems to be much more related to factors other than retinal error. These factors may be related to adaptive changes of central structures that serve learning processes (such as the cerebellum), which may differ from person to person.

Similarly, no correlation was observed between the clinical deficit and the velocity or the duration/amplitude ratio of the nPE. Therefore, in proportion to the saccade amplitude, their duration and velocity show specific adaptive changes in response to the contralateral eye palsy, but these changes do not reflect the extent of the clinical deficit.

### RESULT INTERPRETATION AND COMPARISON WITH THE EXISTING LITERATURE

The amplitude gain of the nPE observed at every optical target distance (5°, 10° and 15°) was generally close to 1. This indicates that the primary saccades were landing accurately on the target so that no significant residual retinal error necessitated corrective (catch-up) saccades. It is known that short-amplitude saccades approach accurately the target, but as the target distance increases, the saccades start to become more hypometric (23–25).

The purpose of the present study was to record the natural adaptive course of the ocular motor system after ANP. Although the examination of larger amplitude saccades (beyond 15°) would provide more data and possibly test the performance limits of the PE more vigorously, we preferred to use saccades up to 15°, as it is proven that this is the true natural range of the saccades in everyday conditions. Gaze displacement beyond 15° is always accompanied by head movements, except in cases where the head is immobilized in the laboratory (26).

Despite the extended literature on saccadic movements in both healthy controls and patients with CNS disease (7,27,28), studies involving paralytic strabismus syndromes are rare. Until the 1970s, examination and follow-up of patients with ocular motor palsy was performed only clinically and, only rarely by the relatively invasive technique of ocular muscle electromyography (29). The gradually increasing availability of electrophysiological methods for measuring ocular movements led to the first studies of ocular palsy.

In 1970, Metz and colleagues (30) recorded saccadic eye movements in two patients with ANP following a head injury as a result of a car accident. DC electroophthalmogram recordings showed that the saccade velocity in the direction of the palsy of the PE was slower than that corresponding to the direction of the healthy (non-paretic) lateral rectus muscle. A few years later, Guntram Kommerell’s team in Freiburg examined two patients with ANP applying the then newly acquired knowledge of phasic and tonic innervation of ocular muscles during saccadic movements (31,32). He concluded that paretic saccades are slower, and assumed that the abrupt antagonist muscle relaxation was important in order to assist the saccade towards the side of the palsy. It should be noted that the recordings were performed with the PE covered.

While reduced velocity of the PE was also noticed in our data, it did not assume statistical significance. It is obvious that the present results cannot be directly compared with the aforementioned studies, not only because of the differences in the recording method, the conditions of the eye coverage and the clinical etiology of the palsy, but also because of the way its angular velocity was calculated. In the present study, taking into account the “main sequence” relation, velocities were presented as normalized fractions of maximal velocity/amplitude, something that was ignored in previous case reports. The same drawback was present in small case series with abducens or oculomotor nerve palsy examined by conventional DC electro-ophthalmography (33,34).

Daroff and colleagues recorded adduction saccades in a patient two weeks after the onset of ocular motor nerve palsy (1). The design of their recordings, although completely different from the present study’s methodology, led to some interesting findings. The researchers permanently covered the nPE and began daily recordings of the PE, which showed gradual improvement of all saccadic dynamics, reaching almost normal values on the sixth day. The eye coverage was then placed on the PE allowing the non-paretic eye to focus on everyday conditions. This led to a rapid, re-deterioration of the PE which within a few days returned to its original state as seen from all motor parameters. That study demonstrated the great potentiality for adaptation of the ocular motor system through adaptive processes within the CNS. In addition to saccadic movements, the vestibulo-ocular reflex has shown similar adaptability, which also improves within a few days by achieving almost normal slow ocular movements after vestibular stimulation on a rotating chair (35). These case reports, however, do not present the natural course of ocular motor recovery, despite forced prolonged use of the PE on a daily basis and therefore any comparison with the present study should be vied with caution. In any case, the finding of Daroff’s team is particularly interesting, due to the fact that the adaptive change of motor parameters during successive recordings did not refer to velocity but the duration and amplitude of the saccades (1). Correspondingly, our data showed that the duration (normalized to the amplitude of the respective saccades) of the nPE was the predominant dynamic parameter which presented the greatest changes, compared with the ocular movements of healthy controls. The PEs, mainly during the second recording session and for the most eccentric saccades (15°), revealed a longer duration of the primary saccade, but similar velocity compared to the eyes of healthy controls.

The saccadic ocular motor system functions ballistically and during the movement is not subject to consciousness. As a result, it is amenable to corrective saccades only after the end of the saccade and after a period at least equal to the system’s latency. This highly automated process is supported by an extensive brainstem neuron network that works seamlessly to align the optical target with the retinal point of the highest sensory neurons concentration. Normal aging, the use of glasses that change the size of the image on the retina and various diseases, such as ANP studied in the present dissertation, however, necessitates a degree of adaptive variability of this network. The cerebellum, with its numerous learning processes, hosts the synapses of climbing fibers to Purkinje cell dendrites (36), and is the predominant candidate structure to undertake such a role. Based on this hypothesis, researchers shifted the ocular muscle tendons to trained monkeys, thus simulating an ANP (37). The animals’ healthy eye was covered in order to accelerate the adaptive adaptation of the saccadic system in the PE. In fact, within 3 days the saccades of the PE regained almost their normal amplitude. In two animals, however, total cerebellectomies were performed and then, after postoperative recovery, the same experimental procedure took place. Unlike animals with intact cerebellum, these monkeys never managed to improve their saccadic dynamics, despite the constant coverage of the HE. This finding proves that the learning process is subject to the saccadic adaptation after a ANP and it takes place in the cerebellum. The same result was reproduced in the study of Vilis and colleagues (12,38), but these researchers argued that the cerebellum is able to change saccadic dynamics disconjugately, thus violating Hering’s law. The latter is also supported by studies showing that experimental damage to deep cerebral nuclei causes unequal asymmetry in both eyes (39), after experimental lesions to the flocculus (40).

Theoretically, two mechanisms can contribute to the the adaptation of the hypometric saccades through a gradual increase in amplitude:

1. parametric change of motor dynamics
2. remapping. The term “map” defines the connection of each point in space to a set of ocular motor commands that can produce a saccade endpoint. With saccadic remapping, a presaccadic increase in neural activity brings an object into a neuron’s receptive field more accurately.

A simple adaptive increase in the saccade amplitude could be congruent with both mechanisms. In the first case, the interaction of the cerebellum with the brainstem saccade generation circuit could affect the amplitude gain of motion. In the second case, a remapping would indicate a change in the visual-sensory representation of space in the brain, or a change in the mapping of space to subsets of motor commands and thus non-motor saccade accuracy improvement. On the contrary, adaptations to other motor dynamics (duration, velocity), in addition to the saccade amplitude, are now compatible with the first adaptation mechanism and not with the remapping process (41). From the findings of the present study, it is obvious that the ANP causes adaptive processes supported by parametric changes in motor dynamics (first mechanism).

Despite the fact that the literature is scarce regarding the saccadic adaptation in ocular nerve or muscle palsy, the adaptation of saccades has been studied in normal populations using the “double step” technique (42,43). This is an experimental example in which the optical target is moved by a computer during a saccade, so that after the end of the saccade, the gaze and the target are not aligned. The remaining retinal error is a parameter, which the CNS adjusts, by gradually changing the amplitude of the primary saccade in order to incorporate this shift from the beginning. Straube and Deubel showed, using the double-step example, that normal subjects who adjusted their saccades to a new, increased amplitude of displacement also increased the normalized duration of saccades (44). This is in line with the results of the present study, that showed increase in the saccade duration of the nPE during the course of the natural adaptive process after a ANP. The same authors emphasize, however, that the adaptive motor changes of the saccadic movements differed considerably from person to person, emphasizing the strong idiosyncratic nature of this adaptive mechanism. This conclusion also emerged in our study, where the size of the initial clinical deficit does not predetermine the magnitude of the saccades’ adaptive changes. The double-step example has been used in many rapid adaptation studies in both humans and non-human primates, most of which showed, in addition to the expected change in the saccade amplitude, and a definite change in other motor parameters, such as velocity, duration and acceleration, but not all studies reach the exact same results (41,45,46). In addition to differences in ocular motion recording and analysis techniques, discrepancies between studies may be due to idiosyncratically individualized adaptation patterns.

Although the above studies were performed in a normal population and were based on an experimental design that can only be used under laboratory conditions (double-step example), they can be compared with our data, which illustrate the adaptation of a pathological process under natural environment conditions. The timing that the adaptation take place is the apparent difference between the two processes: The double-step example induces motor changes rapidly after a few hundred saccades (47), while ANP seems to require at least a few days in order to show signs of motor improvement. However, Scudder and his colleagues conducted experiments on trained monkeys and were able to demonstrate that both rapid adaptation, as achieved by displacing the target in the double-step example, and slower adaptation, such as the one observed after tenectomy or nerve palsy, involve the same neural adaptive mechanism (48). The difference in timing is due to the fact that the adjustment mechanism is closely related to the specific saccade amplitude that the subject is trained at and when the adjustment is completed, this cannot be generalized to the other optical targets amplitudes, for which a similar training must be performed (49,50). Thus, while the double-step example applies to a specific range of saccades in the laboratory, ANP forces the ocular motor system to theoretically adapt infinite amplitudes of saccades as they arise to normal real functional requirements (48). The idiosyncratic differences observed in some of the patients in the present dissertation may be due in part to different ocular motor habits in the participants’ daily activities, that specifically favor the adaptation of some of the optical target amplitudes and disfavor others.

As mentioned above, one of the main findings of the present study is the duration increase of the saccades in the course of adaptive compensation of ANP. This was particularly evident in the second recording session where the nPE performed orthometric saccades but with longer duration than that of healthy controls. Given the fact that the motor command reaches the ocular muscles by a “pulse-step” neural discharge pattern (Figure 5), any adaptive change must be reflected as a change in this pattern.

**Figure 5:**
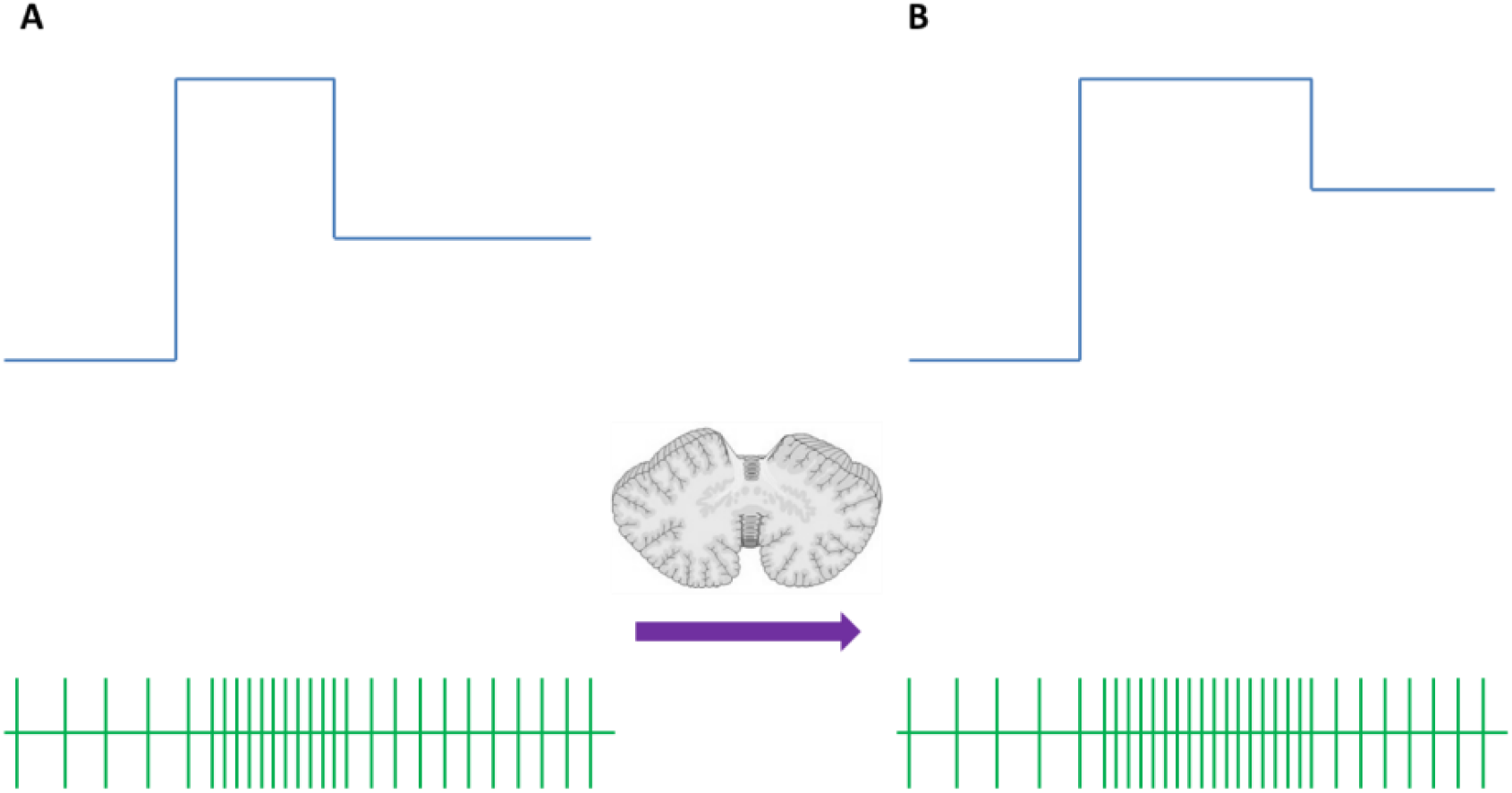
The upper panel schematically depicts the neuron discharge frequency of the abducens nucleus during a saccade. The “pulse-step” pattern is distinct. The lower panel schematically depicts the action potentials, from which the instantaneous discharge frequency of the upper panel is calculated. (A) Discharge pattern before and (B) after saccadic system adaptation. The difference lies in the increase in pulse amplitude.

It has been shown with a similar to the double-step example layout experiment, that normal controls are able to change the pulse of the neural command independently of the step of the same command during saccadic adaptation (51). As the pulse is reflected in the amplitude and velocity of the primary saccade, the step affects the final eye position (which may be that of the primary saccade or is accompanied by corrective secondary saccades or by drift to the target), it is clear that the motor changes we observed relate to the pulse of the motor command. A change in the maximum ocular velocity would be more compatible with an increase of the pulse rate of the neural discharge. However, our results do not show a clear increase in the maximum velocity of the saccades, in either of the recording sessions. The observed increase in duration, on the other hand, indicates an increased amplitude of the pulse motor command (Figure 5). This is consistent with Daroff team’s data of a patient with medial rectus weakness due to oculomotor nerve palsy, where the increased duration of adapted saccades was compatible with the pulse width increment of the respective neural discharge (1). It seems, therefore, that cerebellar learning processes begin immediately after an ocular motor nerve palsy, and in order to increase the range of saccadic movements and achieve accurate saccade endpoints, they enhance the amplitude of the pulse of the neuronal drive. In this way, the eye reaches the target fractions of a second later, but with the right amplitude, virtually zeroing in on retinal error.

The amplitude and width of the pulse are the motor analog of the maximum discharge rate and the discharge duration, respectively, of the saccadic burst generator in the brainstem (52,53). Under normal circumstances, the maximum discharge frequency reaches saturation relatively early, so that when larger amplitude saccades are needed, the burst duration remains the only configurable variable (22). Therefore, the possibility of duration fluctuation of the burst is an already given mode of operation in the healthy ocular motor system, so it is not surprising to use this mechanism from the process of adaptive compensation of ANP, as shown in the present work. Further research could provide more insight into the plastic processes involved in the saccadic adaptation and the understanding of CNS implication to the saccadic disorders.

## Data Availability

All data produced in the present study are available upon reasonable request to the authors

